# Intersectional Disparities in Mental Healthcare Utilization by Sex and Race/Ethnicity among US Adults: An NHANES Study

**DOI:** 10.1101/2025.05.05.25327041

**Authors:** Lotenna Olisaeloka, Gentille Musengimana, Esteban J. Valencia, Daniel Vigo, Ehsan Karim

## Abstract

**Introduction:** Mental healthcare utilization in the US remains low, with sociodemographic factors like sex and race/ethnicity influencing access across different population groups. However, how these factors interact to shape service utilization remains understudied.

**Methods:** Using data from the 2009–2018 cycles of the National Health and Nutrition Examination Survey (NHANES), we employed design-based log-binomial models to estimate the relative differences (prevalence ratios) of mental healthcare utilization across intersecting sex and race/ethnic groups. To assess the absolute differences, we used linear probability regression models to estimate the prevalence differences across these intersectional groups. We also conducted stratified analyses by education, income, health insurance, and depression status, to examine whether disparities persisted across socioeconomic and health-related subgroups.

**Results:** Overall, 9.1% of adults reported accessing mental health services in the past year. Hispanic males had the lowest utilization rates compared to Non-Hispanic (NH) White males, with an adjusted prevalence ratio (aPR) of 0.59 [95% CI: 0.47–0.73]. Among females, significant disparities were observed across all race/ethnic minority groups with NH Black and Hispanic females having significantly lower utilization rates compared to NH White females. Absolute prevalence differences mirrored the relative measures. Stratified analysis were largely consistent with primary results but revealed some variation across education, income and health insurance strata.

**Discussion:** These findings reflect how intersecting sociodemographic characteristics, specifically sex and race/ethnicity, influence utilization of mental health services. Stratified results suggest that socio-economic status may modify these disparities, pointing to the role played by systemic inequities. Hence, culturally informed strategies and wider structural interventions are needed to address these disparities. Future research should consider additional intersecting identities (e.g., sexual orientation and disability) and investigate socio-structural approaches to reducing these gaps.

## INTRODUCTION

Mental health disorders affect approximately 1 in 8 people globally, with prevalence remaining high over the past three decades despite evidence-based treatment options^1–3^. This is partly attributable to a persistent mental health treatment gap, with over 75% of individuals experiencing mental health problems globally not receiving necessary care^4^. In the United States, the number of adults with any mental illness increased by 30% from 2008 to 2019, with over 50 million adults currently affected^5,6^ Yet, more than half of them do not receive care^7,8^. Preventive mental health services like screening and early intervention are also critically lacking, especially for at-risk groups like individuals with chronic physical conditions^9^. Addressing these gaps and ensuring equitable access to mental healthcare presents a pressing public health challenge.

Social determinants of health such as gender, ethnicity, sexual orientation and socioeconomic status influence mental health and access to care^10–12^. Social inequities create barriers to obtaining quality mental healthcare and exacerbate health disparities. Race and ethnicity, in particular are recognized as social constructs shaped by historical, cultural and political factors rather than biological differences. These categories often reflect differential access to resources and opportunities, which influence healthcare access and outcomes^13^. While single-factor disparities by sex and race/ethnicity have been observed, with women and non-Hispanic Whites generally showing higher service utilization rates, the intersection of these factors remain understudied^11,14–18^. Intersectionality, a framework born out of black women’s concurrent experiences of racism and sexism emphasizes the interconnected nature of social inequalities^13,19,20^. The rise of intersectionality in health equity research underscores the limitations of single-factor approaches which treat social categories as independent, and thus fail to capture how the complex interplay of factors shape disparities^19,21^.

Although quantitative methods are crucial for identifying health disparities at the population level, intersectional research has remained largely qualitative due to challenges in translating theoretical constructs into statistical models^19^. This study addresses this gap in relation to mental healthcare utilization by operationalising intersectionality using regression models with interaction terms, as proposed by Bauer (2014)^19^. Specifically, we estimate disparities in mental healthcare utilization among U.S adults across intersections of sex and race/ethnicity. While two axes of identity are not exhaustive, they represent critical starting points for operationalizing intersectionality in healthcare access research^19^. Identifying disproportionately affected intersectional groups can inform targeted interventions and health equity strategies.

## METHODS

### Data Source

We utilized aggregated data from five continuous cycles of the National Health and Nutrition Examination Survey (NHANES) from 2009-2018. The NHANES is an annual population-based, cross-sectional survey of civilian, non-institutionalized U.S. residents conducted by the Centers for Disease Control and Prevention (CDC)^22^. The survey collects sociodemographic, dietary, and health-related information from a nationally representative sample by employing a multi-stage probability sampling methodology. Detailed information on design, methodology and weighting are published on the CDC website. The NHANES receives ethical approval from the CDC Institutional Review Board and all participants gave informed consent^22^. This NHANES dataset is anonymous and publicly available, hence the study is exempt from institutional ethic review as outlined in the institution’s and Tri-Council Policy Statements (TCPS2) on Ethical Conduct for Research Involving Humans^23,24^. We assessed the survey data from 3rd November, 2024 and did not have access to any information that could identify participants at any stage.

### Study Sample

Aggregated data from the five NHANES cycles included 49,693 participants, of which only adults aged 18 years and over were included in the study (n = 30,352). Following the exclusion of individuals with missing values in exposure and outcome, the final analytical sample was 30,340. **Figure 1** illustrates the process of creating the analytical sample.

**Figure 1:**
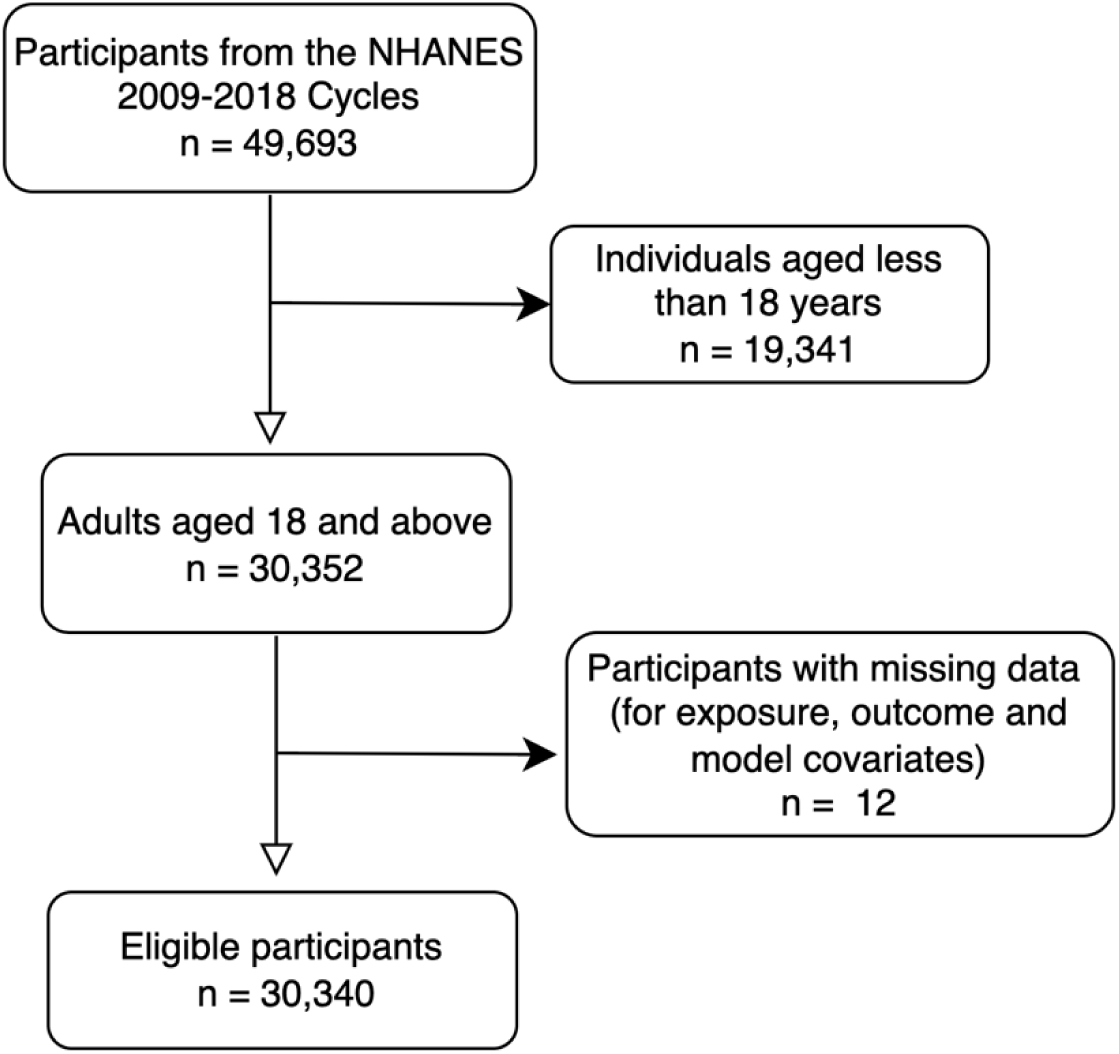
Flowchart illustrating the sample selection process of adults aged 18+ from the NHANES cycles.

### Outcome

The outcome, ‘mental healthcare utilization’ reflected whether respondents accessed mental health services in the past twelve months and was ascertained by an affirmative response to the question: “*During the last 12 months, have you seen or talked to a mental health professional about your health?”* ^25^.

### Exposure

The exposure variables were sex and race/ethnicity as collected by NHANES. Sex was categorized as male or female, and participants were grouped by race/ethnicity variable (RIDRETH1) into four categories (Non-Hispanic White, Hispanic, Non-Hispanic Black and Non-Hispanic Other Race). The ‘Non-Hispanic Other Race’ category included individuals who identified as Non-Hispanic Asian, American Indian or Alaska Native, Native Hawaiian or Other Pacific Islander, and multiracial individuals. This grouping reflects NHANES’ racial and ethnic classifications and was used due to sample size limitations for more granular subgroup analysis.

While race, ethnicity, and sex are immutable characteristics, they serve as proxies for social experiences shaped by systemic factors such as structural racism, discrimination, and privilege. These variables were included to assess disparities in mental healthcare utilization, reflecting the influence of systemic and structural factors.

### Covariate selection

We used a directed acyclic graph (DAG) using *Dagitty* (version 3.1)^26^ to identify suitable risk factors for the outcome which were not positioned along the exposure-outcome causal pathway^27^. Based upon the literature, we considered the following factors: age, education level, marital status, income level, health insurance, perceived health status, depression and chronic physical condition [**Figure 2**]^11,12,15,28–30^. Income level (low, middle, high) was categorized using recommended thresholds of poverty income ratio (PIR)^31^. Perceived health status refers to how participants rated their current general health status (NHANES variable HSD010)^22^. Depression was categorized by severity (none, mild, moderate, moderately severe and severe) based on the Patient Health Questionnaire-9 (PHQ-9) scores^32^. Chronic condition indicated whether participants had diabetes, hypertension, cancer or chronic kidney disease. Age emerged as the sole variable that did not introduce bias when adjusted for [**Figure 2**]. Accordingly, all adjusted models control for age as a precision variable.

**Figure 2:**
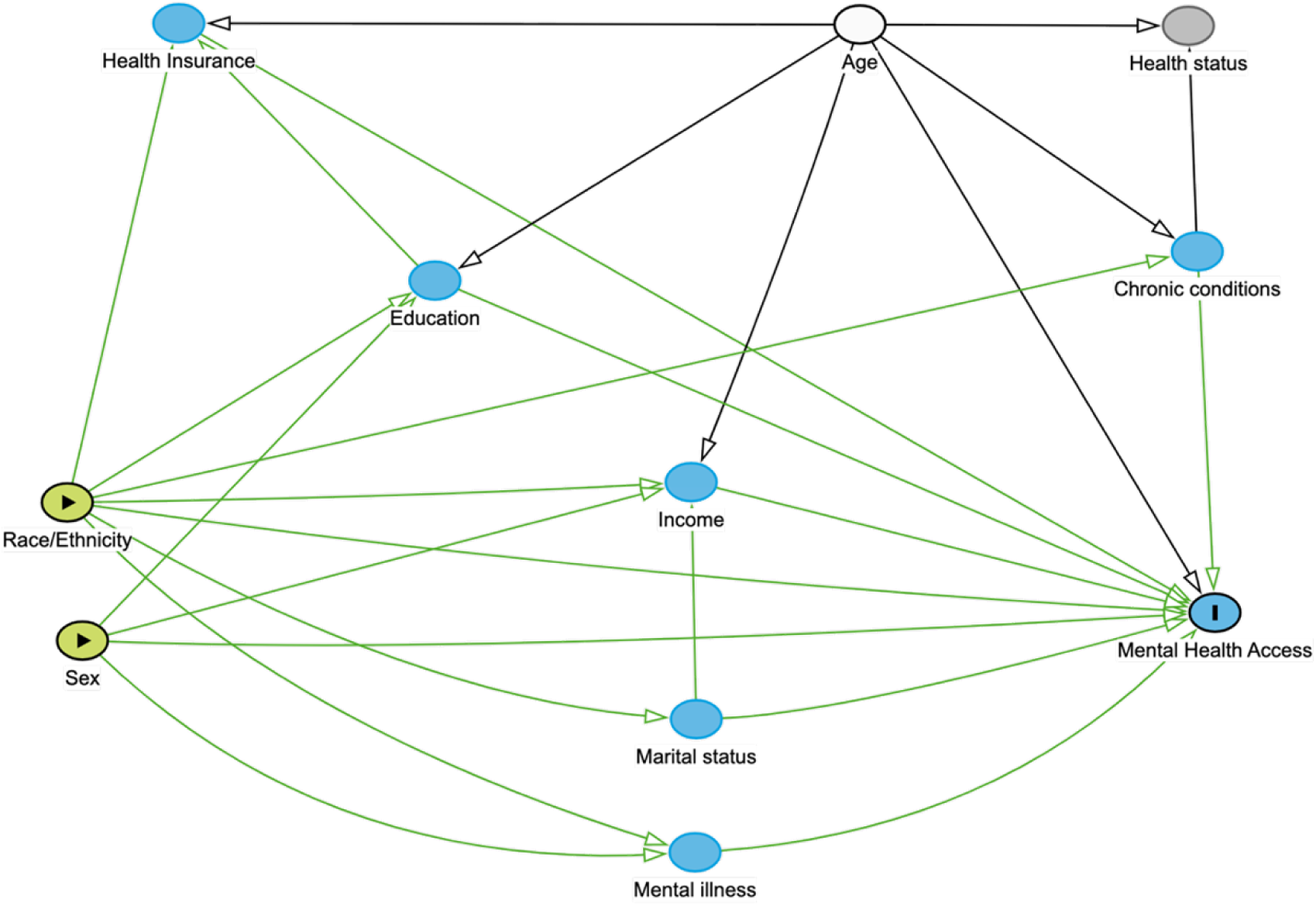
Directed acyclic graph of the relationships between sex, race/ethnicity and mental health service utilization. ***Note:*** Sex and Race/ethnicity are represented as exposures. Mental health access is represented as the outcome. Solid, blue-shaded cells indicate possible mediators of the exposure-outcome relationship. Solid white shaded cells indicate independent risk factors for the outcome. The grey shaded cell indicates a collider variable.

### Statistical Analysis

Descriptive statistics were conducted to summarize the study sample, with sociodemographic and health-related characteristics stratified by sex and race/ethnicity. Categorical variables were presented as counts with their respective weighted percentages. All analyses incorporated survey weights, clusters, and strata to account for the complex survey design^33^.

### Primary Analysis

To operationalize intersectionality in this quantitative analysis, we employed interaction terms between sex and race/ethnicity in our regression models. To examine whether utilization rates differed across intersectional groups, we used design-based log-binomial and linear probability models to estimate interaction on the multiplicative and additive scales respectively^19,34^. Prevalence ratios (PR) and Prevalence Differences (PD), with their 95% confidence intervals (95% CI) and significance levels, were obtained via simple slope/effects analysis. Overall missingness and missingness within variables in the model were less than 5% respectively, hence a complete case analysis was conducted^35^.

### Stratified/Sensitivity Analyses

To assess the consistency of the primary results as well as examine the role of socioeconomic and health-related factors in shaping the observed associations, we conducted stratified analyses by key variables: education level (high school and below vs. college and above), income level (low vs. middle vs. high), and health insurance status (insured vs. uninsured). Stratified analysis was also done by mental illness (depression – yes/no) to assess whether the observed results differed by mental healthcare need. Within each stratum, separate design-based log-binomial regression models including the interaction term between sex and race/ethnicity were fitted. This approach allowed us to extend the intersectional analysis beyond race/ethnicity and sex, evaluating whether the associations and interaction effects observed in the primary analysis differed across socioeconomic groups.

To address missingness in key stratification variables, such as income level (10.5%) and depression (14.2%), we conducted multiple imputation. Little’s MCAR test (p < 0.001) and bivariate analyses respectively revealed that missingness was not completely at random (MCAR) but could be explained by observed data. Under the missing at random (MAR) assumption, our imputation model included variables from the primary model, those requiring imputation (income level, education level, health insurance, and depression), and auxiliary variables (annual family income, marital status, chronic condition, health status, and survey strata). Five imputations with 20 iterations each were performed using logistic regression for binary variables and polynomial regression for categorical variables. The number of iterations was determined by monitoring the trace plots until they demonstrated convergence. Individuals with missing outcome data (n=12) were excluded, resulting in a final analytical sample of 30,340. Design-adjusted multivariable regression models were run on each imputed dataset, and results were pooled using Rubin’s rule which combines the within- and between-imputation variances^36^.

All statistical analyses were conducted using R (v4.3.1). The *survey* package was used to fit the design-adjusted models^37^, and *mice* package was used for missing data analysis. This study followed the Strengthening the Reporting of Observational Studies in Epidemiology (STROBE) reporting guidelines as well as widely adopted recommendations for reporting on sex/gender and race/ethncity^38–40^.

## RESULTS

### Sample Characteristics

The study sample consisted of 30,340 participants, with a nearly even distribution of males (48.5%) and females (51.5%). **Tables 1a and b** present the characteristics of the participants stratified by sex and race/ethnicity respectively. The counts were from the eligible sample while the percentages were derived using survey features, hence represent the national distribution. Significant sex and race-ethnic differences were observed across most demographic and health-related characteristics including education, income level, marital status and health insurance. The sample was predominantly Non-Hispanic (NH) White (64.8%), with small but significant differences in racial/ethnic composition between sexes. Overall, 9.1% of the U.S population accessed mental health services. Females reported a significantly higher proportion of mental health service utilization than males (9.7% vs 8.4%). Stratification by race/ethnicity revealed distinct differences in sociodemographic characteristics and health outcomes. For example, Hispanics had the highest proportion of low-income individuals (30.9%), and NH Whites were significantly more likely to report *excellent* and *very good* health compared to other race/ethnic groups. Mental healthcare utilization was highest among NH Whites (9.8%) and lowest among Hispanic and Other Race groups (7.3% each) [**Table 1b**].

**Table 1a:**
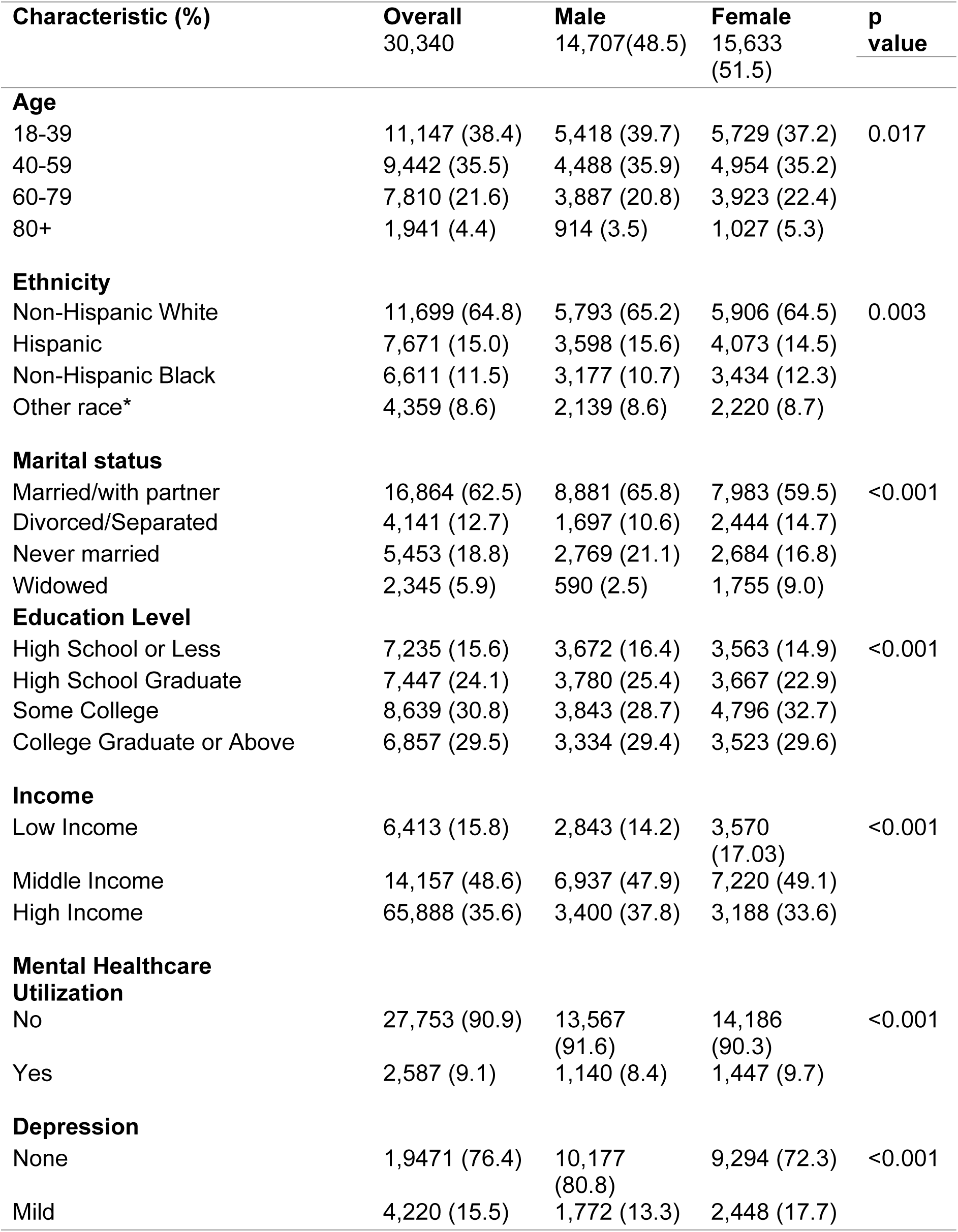
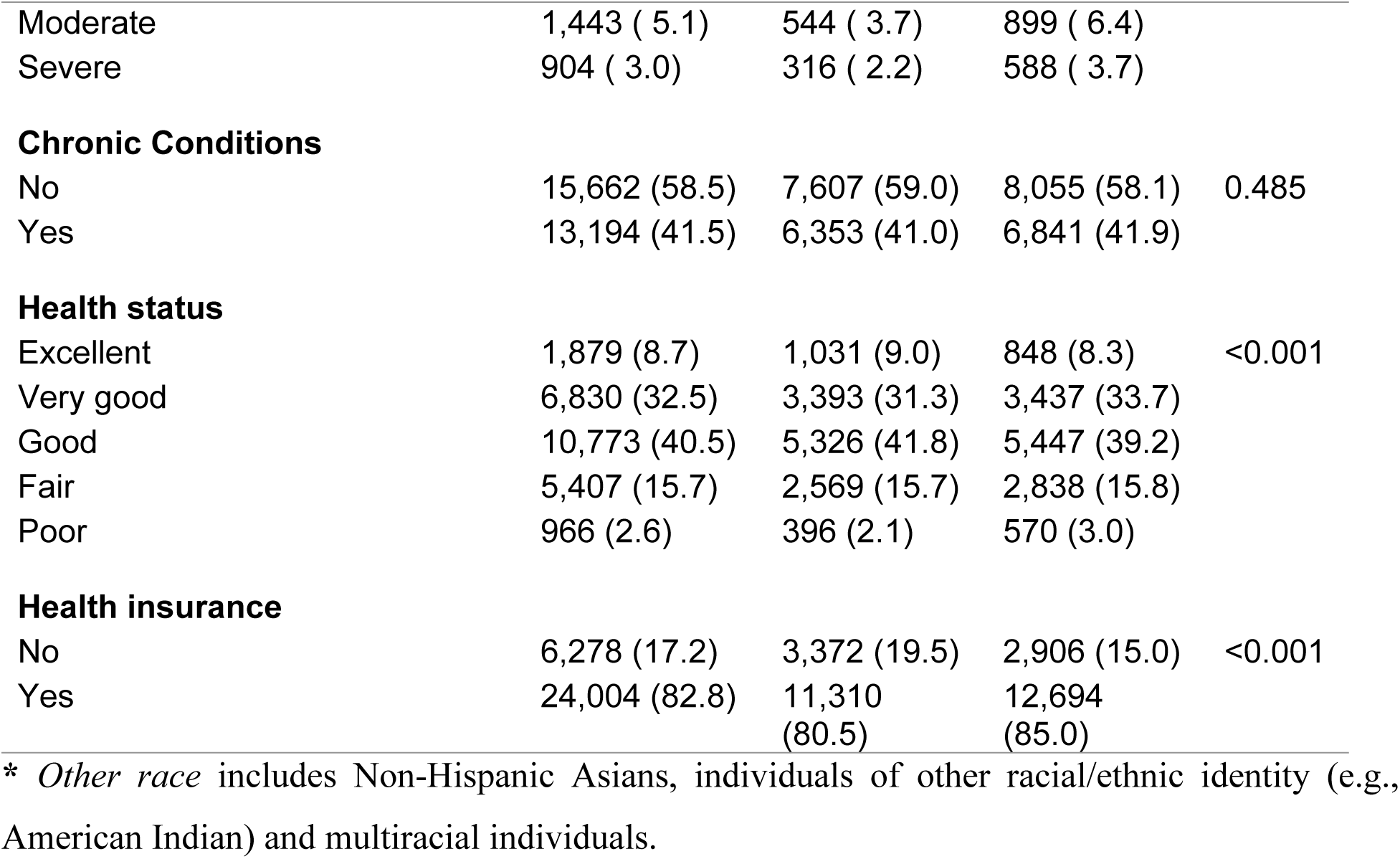
Sample Characteristics by Sex among US adults aged 18 years or more in the 2009-2018 NHANES.

| **Characteristic (%)** | **Overall**<br>**30,340** | **Male**<br>**14,707(48.5)** | **Female**<br>**15,633**<br>**(51.5)** | **p**<br>**value** |
| --- | --- | --- | --- | --- |
| **Age** |  |  |  |  |
| 18-39 | 11,147 (38.4) | 5,418 (39.7) | 5,729 (37.2) | 0.017 |
| 40-59 | 9,442 (35.5) | 4,488 (35.9) | 4,954 (35.2) |  |
| 60-79 | 7,810 (21.6) | 3,887 (20.8) | 3,923 (22.4) |  |
| 80+ | 1,941 (4.4) | 914 (3.5) | 1,027 (5.3) |  |
| **Ethnicity** |  |  |  |  |
| Non-Hispanic White | 11,699 (64.8) | 5,793 (65.2) | 5,906 (64.5) | 0.003 |
| Hispanic | 7,671 (15.0) | 3,598 (15.6) | 4,073 (14.5) |  |
| Non-Hispanic Black | 6,611 (11.5) | 3,177 (10.7) | 3,434 (12.3) |  |
| Other race* | 4,359 (8.6) | 2,139 (8.6) | 2,220 (8.7) |  |
| **Marital status** |  |  |  |  |
| Married/with partner | 16,864 (62.5) | 8,881 (65.8) | 7,983 (59.5) | <0.001 |
| Divorced/Separated | 4,141 (12.7) | 1,697 (10.6) | 2,444 (14.7) |  |
| Never married | 5,453 (18.8) | 2,769 (21.1) | 2,684 (16.8) |  |
| Widowed | 2,345 (5.9) | 590 (2.5) | 1,755 (9.0) |  |
| **Education Level** |  |  |  |  |
| High School or Less | 7,235 (15.6) | 3,672 (16.4) | 3,563 (14.9) | <0.001 |
| High School Graduate | 7,447 (24.1) | 3,780 (25.4) | 3,667 (22.9) |  |
| Some College | 8,639 (30.8) | 3,843 (28.7) | 4,796 (32.7) |  |
| College Graduate or Above | 6,857 (29.5) | 3,334 (29.4) | 3,523 (29.6) |  |
| **Income** |  |  |  |  |
| Low Income | 6,413 (15.8) | 2,843 (14.2) | 3,570<br>(17.03) | <0.001 |
| Middle Income | 14,157 (48.6) | 6,937 (47.9) | 7,220 (49.1) |  |
| High Income | 65,888 (35.6) | 3,400 (37.8) | 3,188 (33.6) |  |
| **Mental Healthcare**<br>**Utilization** |  |  |  |  |
| No | 27,753 (90.9) | 13,567<br>(91.6) | 14,186<br>(90.3) | <0.001 |
| Yes | 2,587 (9.1) | 1,140 (8.4) | 1,447 (9.7) |  |
| **Depression** |  |  |  |  |
| None | 1,9471 (76.4) | 10,177<br>(80.8) | 9,294 (72.3) | <0.001 |
| Mild | 4,220 (15.5) | 1,772 (13.3) | 2,448 (17.7) |  |
| Moderate | 1,443 ( 5.1) | 544 ( 3.7) | 899 ( 6.4) |  |
| Severe | 904 ( 3.0) | 316 ( 2.2) | 588 ( 3.7) |  |
| **Chronic Conditions** |  |  |  |  |
| No | 15,662 (58.5) | 7,607 (59.0) | 8,055 (58.1) | 0.485 |
| Yes | 13,194 (41.5) | 6,353 (41.0) | 6,841 (41.9) |  |
| **Health status** |  |  |  |  |
| Excellent | 1,879 (8.7) | 1,031 (9.0) | 848 (8.3) | <0.001 |
| Very good | 6,830 (32.5) | 3,393 (31.3) | 3,437 (33.7) |  |
| Good | 10,773 (40.5) | 5,326 (41.8) | 5,447 (39.2) |  |
| Fair | 5,407 (15.7) | 2,569 (15.7) | 2,838 (15.8) |  |
| Poor | 966 (2.6) | 396 (2.1) | 570 (3.0) |  |
| **Health insurance** |  |  |  |  |
| No | 6,278 (17.2) | 3,372 (19.5) | 2,906 (15.0) | <0.001 |
| Yes | 24,004 (82.8) | 11,310 (80.5) | 12,694 (85.0) |  |

\* *Other race* includes Non-Hispanic Asians, individuals of other racial/ethnic identity (e.g., American Indian) and multiracial individuals.

**Table 1b.**
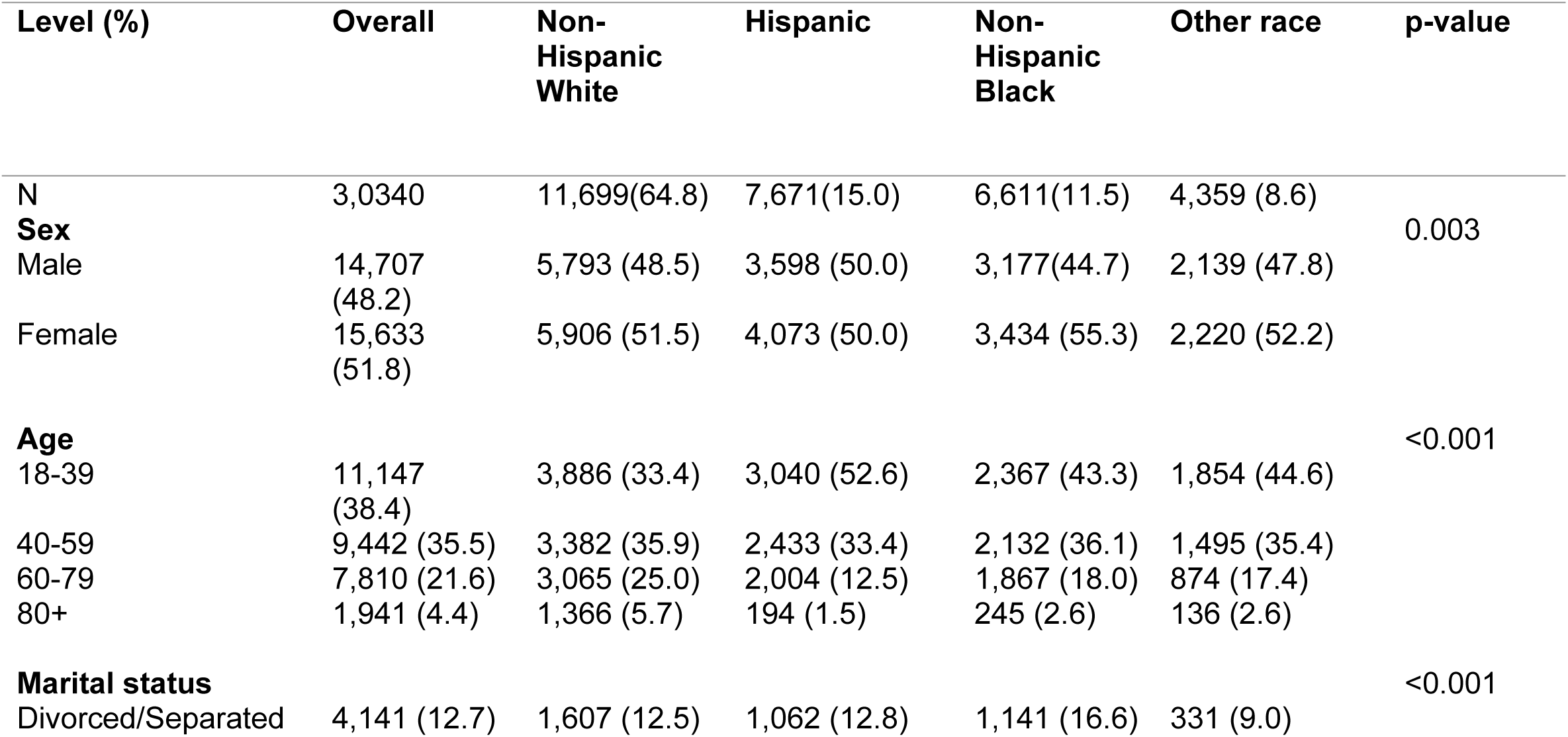
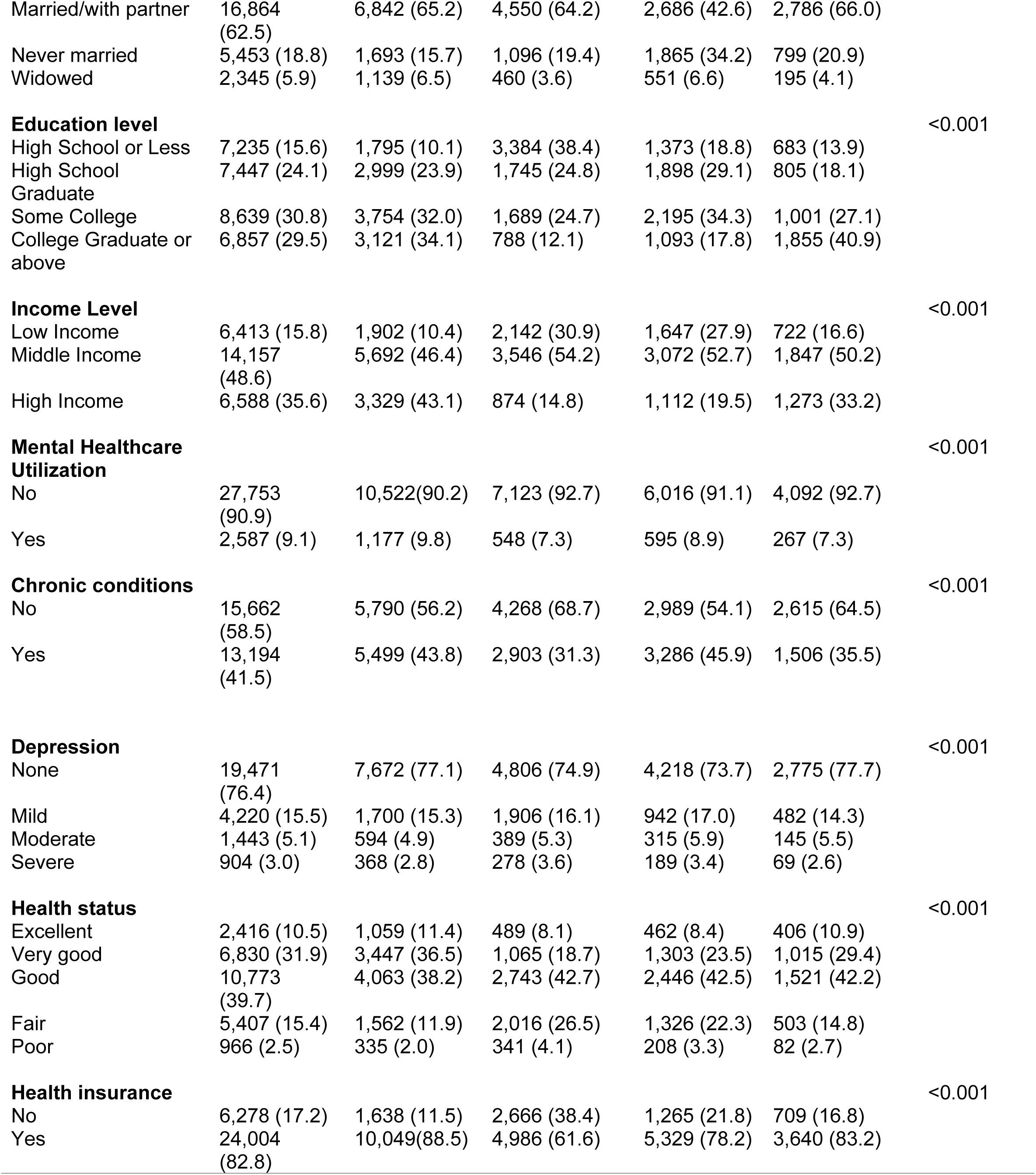
Sample Characteristics stratified by Race-ethnicity among US adults aged 18 years or more in the 2009-2018 NHANES.

| Level (%) | Overall | Non-Hispanic White | Hispanic | Non-Hispanic Black | Other race | p-value |
| --- | --- | --- | --- | --- | --- | --- |
| **N** | 3,0340 | 11,699(64.8) | 7,671(15.0) | 6,611(11.5) | 4,359 (8.6) |  |
| **Sex** |  |  |  |  |  | 0.003 |
| Male | 14,707 (48.2) | 5,793 (48.5) | 3,598 (50.0) | 3,177(44.7) | 2,139 (47.8) |  |
| Female | 15,633 (51.8) | 5,906 (51.5) | 4,073 (50.0) | 3,434 (55.3) | 2,220 (52.2) |  |
| **Age** |  |  |  |  |  | <0.001 |
| 18-39 | 11,147 (38.4) | 3,886 (33.4) | 3,040 (52.6) | 2,367 (43.3) | 1,854 (44.6) |  |
| 40-59 | 9,442 (35.5) | 3,382 (35.9) | 2,433 (33.4) | 2,132 (36.1) | 1,495 (35.4) |  |
| 60-79 | 7,810 (21.6) | 3,065 (25.0) | 2,004 (12.5) | 1,867 (18.0) | 874 (17.4) |  |
| 80+ | 1,941 (4.4) | 1,366 (5.7) | 194 (1.5) | 245 (2.6) | 136 (2.6) |  |
| **Marital status** |  |  |  |  |  | <0.001 |
| Divorced/Separated | 4,141 (12.7) | 1,607 (12.5) | 1,062 (12.8) | 1,141 (16.6) | 331 (9.0) |  |
| Married/with partner | 16,864<br>(62.5) | 6,842 (65.2) | 4,550 (64.2) | 2,686 (42.6) | 2,786 (66.0) |  |
| Never married | 5,453 (18.8) | 1,693 (15.7) | 1,096 (19.4) | 1,865 (34.2) | 799 (20.9) |  |
| Widowed | 2,345 (5.9) | 1,139 (6.5) | 460 (3.6) | 551 (6.6) | 195 (4.1) |  |
| **Education level** |  |  |  |  |  | <0.001 |
| High School or Less | 7,235 (15.6) | 1,795 (10.1) | 3,384 (38.4) | 1,373 (18.8) | 683 (13.9) |  |
| High School Graduate | 7,447 (24.1) | 2,999 (23.9) | 1,745 (24.8) | 1,898 (29.1) | 805 (18.1) |  |
| Some College | 8,639 (30.8) | 3,754 (32.0) | 1,689 (24.7) | 2,195 (34.3) | 1,001 (27.1) |  |
| College Graduate or above | 6,857 (29.5) | 3,121 (34.1) | 788 (12.1) | 1,093 (17.8) | 1,855 (40.9) |  |
| **Income Level** |  |  |  |  |  | <0.001 |
| Low Income | 6,413 (15.8) | 1,902 (10.4) | 2,142 (30.9) | 1,647 (27.9) | 722 (16.6) |  |
| Middle Income | 14,157<br>(48.6) | 5,692 (46.4) | 3,546 (54.2) | 3,072 (52.7) | 1,847 (50.2) |  |
| High Income | 6,588 (35.6) | 3,329 (43.1) | 874 (14.8) | 1,112 (19.5) | 1,273 (33.2) |  |
| **Mental Healthcare Utilization** |  |  |  |  |  | <0.001 |
| No | 27,753<br>(90.9) | 10,522(90.2) | 7,123 (92.7) | 6,016 (91.1) | 4,092 (92.7) |  |
| Yes | 2,587 (9.1) | 1,177 (9.8) | 548 (7.3) | 595 (8.9) | 267 (7.3) |  |
| **Chronic conditions** |  |  |  |  |  | <0.001 |
| No | 15,662<br>(58.5) | 5,790 (56.2) | 4,268 (68.7) | 2,989 (54.1) | 2,615 (64.5) |  |
| Yes | 13,194<br>(41.5) | 5,499 (43.8) | 2,903 (31.3) | 3,286 (45.9) | 1,506 (35.5) |  |
| **Depression** |  |  |  |  |  | <0.001 |
| None | 19,471<br>(76.4) | 7,672 (77.1) | 4,806 (74.9) | 4,218 (73.7) | 2,775 (77.7) |  |
| Mild | 4,220 (15.5) | 1,700 (15.3) | 1,906 (16.1) | 942 (17.0) | 482 (14.3) |  |
| Moderate | 1,443 (5.1) | 594 (4.9) | 389 (5.3) | 315 (5.9) | 145 (5.5) |  |
| Severe | 904 (3.0) | 368 (2.8) | 278 (3.6) | 189 (3.4) | 69 (2.6) |  |
| **Health status** |  |  |  |  |  | <0.001 |
| Excellent | 2,416 (10.5) | 1,059 (11.4) | 489 (8.1) | 462 (8.4) | 406 (10.9) |  |
| Very good | 6,830 (31.9) | 3,447 (36.5) | 1,065 (18.7) | 1,303 (23.5) | 1,015 (29.4) |  |
| Good | 10,773<br>(39.7) | 4,063 (38.2) | 2,743 (42.7) | 2,446 (42.5) | 1,521 (42.2) |  |
| Fair | 5,407 (15.4) | 1,562 (11.9) | 2,016 (26.5) | 1,326 (22.3) | 503 (14.8) |  |
| Poor | 966 (2.5) | 335 (2.0) | 341 (4.1) | 208 (3.3) | 82 (2.7) |  |
| **Health insurance** |  |  |  |  |  | <0.001 |
| No | 6,278 (17.2) | 1,638 (11.5) | 2,666 (38.4) | 1,265 (21.8) | 709 (16.8) |  |
| Yes | 24,004<br>(82.8) | 10,049(88.5) | 4,986 (61.6) | 5,329 (78.2) | 3,640 (83.2) |  |

### Mental Healthcare Utilization across Intersectional Groups

We observed significant disparities in relative and absolute prevalence estimates of mental healthcare utilization across sex and race/ethnic intersectional groups **[Table 2].** Compared to NH White males, Hispanic males were the least likely (41% less likely, aPR = 0.59 [0.47–0.73]) to utilize services, followed by males in the Other race group (33% less likely, aPR = 0.66 [0.50– 0.88]). NH Black males were also less likely to utilize mental health services compared to NH White males. However, this association was insignificant (aPR = 0.91 [0.75–1.12]). Among females, Hispanic, NH Blacks and women in the Other race category were significantly less likely (25%, 19% and 27% less likely respectively) to utilize mental health services compared to NH White females.

**Table 2:** Prevalence Ratios (PR) and Prevalence Differences (PD) of Mental Healthcare Utilization by Sex and Race/Ethnicity (95% Confidence Intervals)

| Race/Ethnicity<br>PR & PD (95% CI) | Male | Female | Female vs Male<br>(within strata of<br>race-ethnicity) |
| --- | --- | --- | --- |
| **Relative Scale** |  |  |  |
| **Unadjusted PR** |  |  |  |
| Non-Hispanic White | Ref<sup>^</sup> | Ref | 1.14 (0.97, 1.32) |
| Hispanic | 0.65 (0.52, 0.80)*** | 0.84 (0.70, 0.99)* | 1.46 (1.20, 1.79)*** |
| Non-Hispanic Black | 0.97 (0.80, 1.19) | 0.87 (0.73, 1.05) | 1.02 (0.87, 1.20) |
| Other Race | 0.70 (0.53, 0.92)** | 0.79 (0.61, 1.00)* | 1.27 (0.92, 1.75) |
| **Adjusted PR** |  |  |  |
| Non-Hispanic White | Ref | Ref | 1.16 (1.00, 1.36)* |
| Hispanic | 0.59 (0.47, 0.73)*** | 0.75 (0.63, 0.90)*** | 1.49 (1.22, 1.80)** |
| Non-Hispanic Black | 0.91 (0.75, 1.12) | 0.81 (0.68, 0.98)* | 1.04 (0.89, 1.21) |
| Other Race | 0.66 (0.50, 0.88)** | 0.73 (0.57, 0.92)* | 1.27 (0.92, 1.77) |
| **Absolute Scale** |  |  |  |
| **Unadjusted PD** |  |  |  |
| Non-Hispanic White | Ref | Ref | 0.01 (-0.00, 0.03) |
| Hispanic | -0.03 (-0.05, -0.02)*** | -0.02 (-0.03, -0.00)* | 0.03 (0.01, 0.04)*** |
| Non-Hispanic Black | -0.00 (-0.02, 0.02) | -0.01 (-0.03, 0.00) | 0.00 (-0.01, 0.02) |
| Other Race | -0.03 (-0.05, -0.01)** | -0.02 (-0.04, -0.00)* | 0.02 (-0.01, 0.04) |
| **Adjusted PD** |  |  |  |
| Non-Hispanic White | Ref | Ref | 0.01 (-0.00, 0.03) |
| Hispanic | -0.03 (-0.05, -0.01)*** | -0.02 (-0.03, -0.00)** | 0.03 (0.01, 0.04)*** |
| Non-Hispanic Black | -0.00 (-0.02, 0.01) | -0.02 (-0.03, 0.00) | -0.00 (-0.01, 0.01) |
| Other Race | -0.03 (-0.05, -0.01)** | -0.02 (-0.04, -0.00)* | 0.02 (-0.01, 0.04) |

Adjusted estimates are adjusted for Age.
^Ref are the reference groups of comparisons. Within male and female categories, individuals in other intersectional groups are compared to Non-Hispanic Whites.
\*indicates statistical significance at p values: \*<0.05, \*\*<0.01 \*\*\*<0.001

Within the same race-ethnic group, females were more likely to utilize services than males. While NH White (aPR = 1.16 [1.00, 1.36]) and Hispanic females (aPR = 1.49 [1.22, 1.80]) had significantly higher probabilities of utilizing services than their male counterparts, this association was not significant among NH Blacks and individuals in the Other race category.

Results on the absolute/additive scale were consistent with the relative findings but indicated smaller differences [**Table 2**].

### Stratified Analysis

The results of the stratified analyses were largely consistent with the primary analysis, although few subgroup differences emerged across education, income, health insurance, and depression status. [**Table 3**].

**Table 3:** Results of Stratified Analysis by Education level, Income level, Health Insurance and Depression status showing adjusted Prevalence Ratios (PR) with 95% CIs.

| Stratification Variable | Male<br>aPR (95% CI) | Female<br>aPR (95% CI) | Female vs Male<br>aPR (95% CI) |
| --- | --- | --- | --- |
| **Education** |  |  |  |
| **High School or below** |  |  |  |
| Non-Hispanic White | Ref | Ref | 1.17 [0.91; 1.51] |
| Hispanic | 0.58 [0.44,0.75]*** | 0.74 [0.58,0.93]** | 1.51 [1.17,1.92]*** |
| Non-Hispanic Black | 1.26 [0.96, 1.65] | 0.93 [0.72, 1.21] | 0.87 [0.68; 1.12] |
| Other race | 0.89 [0.59, 1.35] | 0.94 [0.64, 1.38] | 1.25 [0.73, 2.12] |
| **College and above** |  |  |  |
| Non-Hispanic White | Ref | Ref | 1.14[0.96, 1.35] |
| Hispanic | 0.73 [0.54,0.99]* | 0.88[0.69,1.12] | 1.38[1.02; 1.88]* |
| Non-Hispanic Black | 0.70 [0.55, 0.88]** | 0.78[0.63, 0.97]* | 1.28[0.99; 1.67] |
| Other race | 0.58[0.39, 0.86]** | 0.66 [0.50, 0.87]** | 1.31[0.83, 2.06] |
| **Income** |  |  |  |
| **Low Income** |  |  |  |
| Non-Hispanic White | Ref | Ref | 1.22 [0.99, 1.51] |
| Hispanic | 0.46 [0.36, 0.60]*** | 0.47[0.36, 0.62]*** | 1.01 [0.73, 1.39] |
| Non-Hispanic Black | 1.01 [0.77, 1.31] | 0.67 [0.50, 0.99]** | 0.72 [0.52, 0.99] |
| Other race | 0.68 [0.43, 1.10] | 0.62 [0.44, 0.90]** | 1.06 [0.64, 1.75] |
| **Middle Income** |  |  |  |
| Non-Hispanic White | Ref | Ref | 1.15 [0.89, 1.49] |
| Hispanic | 0.56 [0.40, 0.79]*** | 0.75[0.46, 1.06] | 1.62 [1.20, 2.18]** |
| Non-Hispanic Black | 0.75 [0.55, 1.03] | 0.75 [0.52, 1.57] | 1.26 [0.93, 1.72] |
| Other race | 0.45[0.28, 0.75]*** | 0.57 [0.29, 1.14] | 1.65 [0.95, 2.82] |
| **High Income** |  |  |  |
| Non-Hispanic White | Ref | Ref | 1.11 [0.84, 0.46] |
| Hispanic | 0.73 [0.46, 1.15] | 1.00[0.63, 0.90] | 1.54 [0.89, 2.67] |
| Non-Hispanic Black | 0.72 [0.49, 1.06] | 0.95 [0.57, 1.57] | 0.95 [0.57, 1.57] |
| Other race | 0.76 [0.43, 1.36] | 0.89 [0.44, 1.73] | 0.87 [0.44, 1.73] |
| **Insurance** |  |  |  |
| **Uninsured** |  |  |  |
| Non-Hispanic White | Ref | Ref | 1.75[1.21; 2.53]** |
| Hispanic | 0.52 [0.34,0.81]** | 0.30 [0.21,0.41]*** | 1.00 [0.73,1.37] |
| Non-Hispanic Black | 0.87[0.54, 1.37] | 0.45[0.30,0.70]*** | 0.92[0.59; 1.43] |
| Other race | 0.73 [0.33, 1.62] | 0.43[0.25, 0.76]** | 1.04[0.47, 2.32] |
| **Insured** |  |  |  |
| Non-Hispanic White | Ref | Ref | 1.11[0.94, 1.28] |
| Hispanic | 0.73 [0.57,0.94]* | 1.00[0.84,1.20] | 1.51[1.20; 1.92]*** |
| Non-Hispanic Black | 0.99[0.80, 1.12] | 0.90[0.74, 1.08] | 1.00[0.86; 1.17] |
| Other race | 0.67[0.49, 0.90]** | 0.78[0.79, 1.01] | 1.28[0.89, 1.84] |
| **Depression status** |  |  |  |
| **No Depression** |  |  |  |
| Non-Hispanic White | Ref | Ref | 1.08[0.86; 1.37] |
| Hispanic | 0.55[0.42,0.72]*** | 0.75[0.57,0.98]* | 1.48[1.13,1.91]** |
| Non-Hispanic Black | 0.88[0.66, 1.17] | 0.73[0.52, 0.90]** | 0.84[0.66; 1.09] |
| Other race | 0.57 [0.34, 0.93]* | 0.73 [0.52, 1.04] | 1.40[0.80, 2.46] |
| **Depression** |  |  |  |
| Non-Hispanic White | Ref | Ref | 0.99[0.81, 1.21] |
| Hispanic | 0.61[0.46,0.81]*** | 0.70[0.57,0.87]*** | 1.14[0.89; 1.46] |
| Non-Hispanic Black | 0.90[0.70, 1.15] | 0.84[0.68, 1.03] | 0.92[0.76; 1.12] |
| Other race | 0.82[0.57, 1.17] | 0.76 [0.57, 1.01] | 0.91[0.62, 1.37] |

### Education

Compared to the primary analysis, stratification by education revealed notable differences among NH Black individuals. Among those with *high school education or less*, NH Black males had higher mental health service utilization than NH White males (aPR = 1.26, 95% CI: 0.96–1.65), contrasting with the overall finding of lower utilization among NH Black males in the primary analysis. NH Black males with *high school education or less* were also more likely to utilize services compared to their female counterparts (NH Black females) [**Table 3**]. Although these differences were not statistically significant, they indicate a directional reversal in disparities among lower-educated NH Black individuals compared to results from the primary analysis. Conversely, among those with higher education (*college and above*), findings mirror the main analysis with NH Black males having significantly lower utilization than NH White males, and NH Black females having higher utilization than NH Black males.

### Income

Among individuals in the *middle-income* category, results from the stratified analysis aligned closely with the overall findings. However, among those with *low income*, NH Black females had lower utilization than NH Black males (aPR = 0.72 [0.52, 0.99]), again reversing the sex pattern from the primary analysis. This reversal is consistent with the pattern seen in the lower education stratum, suggesting a potential intersection of sex and socio-economic status in shaping service use. Among individuals with high-income, disparities across intersectional groups were not statistically significant. This contrasts with the significant disparities seen in the main analysis, suggesting that higher income may attenuate disparities.

### Insurance

Stratified results by insurance status were broadly consistent with the primary analysis, particularly in terms of direction. However, disparities were more pronounced among uninsured females, with ethnic minority females having even lower utilization rates than in the main analysis. For example, uninsured Hispanic women had 70% lower utilization than uninsured NH White women (aPR = 0.30, 95% CI: 0.21–0.41), a sharper disparity than observed in the primary analysis (25% lower utilization, aPR = 0.75), indicating that lack of insurance may exacerbate disparities among women.

### Depression

The adjusted prevalence ratios (aPRs) for mental healthcare utilization stratified by depression status were largely consistent with the primary analysis suggesting that depression status did not substantially alter the observed disparities.

## Discussion

### Main Findings

This study examined disparities in mental healthcare utilization among U.S adults across intersections of sex and race-ethnicity using data from NHANES 2009-2018 cycles. Overall, only 9.1% of adults in the United States reported accessing mental health services in the past year, which aligns with existing research documenting low utilization rates despite high prevalence of mental health needs^7,8^. The primary interaction analysis demonstrated that mental healthcare utilization varied significantly across intersecting sex and race/ethnicity groups. Among males, Hispanic men were the least likely (41% less likely) to access mental healthcare relative to NH White men. Similarly, females across all minority race/ethnicity groups were significantly less likely to access services than NH White females, with prevalence ratios ranging from 0.75 to 0.81. Within the same racial/ethnic groups, females generally accessed services more than their male counterparts. Prevalence estimates on the absolute scale aligned with findings on the relative scale although the absolute differences were smaller which possibly reflects the low overall prevalence of mental healthcare utilization in the sample.

Study findings generally align with existing literature that show females having high utilization rates than males and NH whites having higher rates than other race/ethnic groups. However, by applying an intersectional framework and modelling prevalence estimates on relative and absolute scales, this analysis captured nuanced disparities that might otherwise remain obscured by single factor approaches^11,17,21^. For instance, Hispanic males demonstrated the lowest utilization rates, representing a crucial disparity point requiring targeted intervention^41^. Further, the varying magnitude of sex gaps across race-ethnic groups—from 49% higher rates for Hispanic females to only 4% for NH Black females compared to their male counterparts—suggests that sex and race-ethnicity interact differently across groups.

The observed disparity patterns likely reflect complex interactions between social norms, help-seeking behaviors, and systemic factors rather than differences in mental health diagnosis or needs. Research shows that minority ethnic groups like Hispanics and NH Blacks have similar or higher prevalence of mental disorders compared to NH Whites^18,42,43^. This is also reflected in this study sample in which Hispanics and NH Blacks have higher prevalence of depression severity than NH Whites. Further, the prevalence ratios for mental healthcare utilization across depression strata were largely consistent with the main analysis, suggesting that disparities in access persist regardless of mental health needs. This finding aligns with Crenshaw’s intersectionality theory and existing research on how intersecting social identities, rather than clinical need alone, influence healthcare access and utilization^18,20,30^.

Several factors might contribute to these observed intersectional disparities. Cultural and systemic barriers play significant roles, including limited cultural competence among healthcare providers, varying cultural conceptualizations of mental health and illness, and differential stigma across cultural groups^44,45^. Additionally, structural barriers such as economic disparities, and insurance coverage gaps likely contribute to these patterns^5^. For instance, the pronounced disparities among Hispanic males might reflect the intersection of traditional masculine norms alongside barriers disproportionately affecting Hispanics including language barriers and immigration-related issues such as being undocumented, working low-income jobs, and not having health insurance^41,44,46–50^.

### Stratified Analysis

In the stratified analysis, some differences observed among some subgroups highlight how socioeconomic factors might modulate mental health access disparities. For example, lack of health insurance amplified disparities across sex and race/ethnicity, particularly for Hispanic and NH Black females. Likewise, income level seemingly influenced mental healthcare utilization, particularly for NH Blacks and Hispanics. Additionally, the absence of significant differences among high-income individuals suggests that socioeconomic advantages may reduce disparities observed in the overall sample.

Education also appeared to influence mental healthcare utilization rates particularly among NH Blacks. In contrast to the primary results and findings from individuals in the higher education category, NH Black males with low education had higher utilization rates than NH White males. Additionally, NH Black females in this group were significantly less likely to access services than NH Black males. This suggests that the overall non-significant difference in utilization observed for NH Black males compared to NH White males and NH Black females may be due to the higher service utilization among NH Black males with lower education. This finding possibly reflects diagnostic patterns, as Black men, particularly those with lower education and socioeconomic status are more likely than White men and Black women to be diagnosed with severe mental illness and substance use disorder^42,45^. Consequently, they are more likely to enter treatment through involuntary or mandated pathways including criminal justice referrals and emergency services rather than voluntary help-seeking^42,45,51^.

### Policy and Practice Implications

This study’s findings have important implications for public health practice and policy. They suggest the need to better understand reasons for the observed disparities and for targeted multi-level interventions that address both socio-cultural and structural barriers to mental health service utilization. At the healthcare system level, increasing diversity and cultural competency of mental healthcare professionals, and implementing culturally informed mental health interventions could address the unique needs and preferences of different intersectional groups, and help reduce disparities. Community-level interventions might include engaging community leaders in mental health outreach, developing culturally specific anti-stigma campaigns, and creating targeted programs for underserved intersectional groups, particularly Hispanic males^28,42,50,52^. Wider structural reforms are also essential to address socioeconomic barriers to mental healthcare. Expanding Medicaid and other insurance programs can reduce cost-related access barriers, while mobile clinics and telehealth programs, can help reach individuals in underserved areas^5,52,53^. Policies addressing social determinants of health including education, employment, and housing are also critical for achieving equitable access to care^52,54^. Addressing these intersectional disparities is critical not only for improving population mental health outcomes but also for broader public health and socioeconomic benefits, including enhanced productivity and increased quality of life^55^.

### Limitations

Several limitations should be considered when interpreting these findings. First, NHANES’ use of binary sex assigned at birth rather than gender identity represents a significant limitation^25^. While sex and gender are correlated, they are distinct constructs. Thus, this limitation masks important nuances in mental health service utilization among gender-diverse populations. This is particularly important as research shows that transgender and non-binary individuals experience disproportionately higher rates of mental health challenges and face unique barriers to care^56^.

Second, intersectionality is not limited to sex or gender and race/ethnicity but also include other characteristics like sexual orientation which could not be included in this analysis despite being important given well-documented mental health disparities among LGBTQ+ populations^21,56^. This limitation stemmed from methodological challenges in the NHANES dataset including insufficient sample sizes for non-heterosexual groups even after combining multiple cycles and high rates of non-specific responses to sexual orientation questions (e.g., ‘don’t know’, ‘refused’, ‘not sure’ or ‘something else’). Thus, the National Centre for Health Statistics discourages the use of this variable for analysis^22^. This reflects a broader challenge in population health data systems, highlighting the need for improved data collection methods that better capture sexual orientation while maintaining statistical power for meaningful analysis. Future research should prioritize the inclusion of more comprehensive sociodemographic variables to capture the complexity of intersecting identities. Surveys and data collection tools could be enhanced by integrating better measures for gender identity, sexual orientation and disability.

Additional limitations include reliance on self-reported service utilization, which may be subject to recall and social desirability bias. Further, the exclusion of institutionalized populations from NHANES may miss important segments of the population needing mental health care. Despite these inherent limitations, this study leveraged an intersectional approach and utilized a large nationally representative sample to explore disparities in mental healthcare utilization across intersections of sex and race/ethnicity with additional consideration of other axes via stratified analysis.

## Conclusion

Our findings reveal significant intersectional disparities in mental health service utilization in the United States, particularly by sex and race/ethnicity. These disparities are further shaped by socioeconomic factors such as education and income. These results underscore the need to consider multiple, intersecting identities when designing policies and interventions to improve mental healthcare access. Addressing these layered inequities will require both culturally responsive approaches and structural reforms to reduce barriers and promote equitable access to care.

## Data Availability

The National Health and Nutrition Survey (NHANES) dataset used for this study is publicly available via the CDC website.

## Authors Contribution(s)

L.O. conceptualized the study, developed the methodology, conducted the formal analysis, curated the data, wrote the original draft and revised the manuscript. G.M. contributed to the methodology, analysis and drafting of the manuscript. E.V. and E.K. assisted with analysis, writing, editing, and revision. D.V. contributed to revision and supervision. All authors reviewed and approved the final manuscript.

